# Parkinson’s Disease Recognition using a Gamified Website: Machine Learning Feasibility Study

**DOI:** 10.1101/2023.08.22.23294440

**Authors:** Shubham Parab, Jerry R Boster, Peter Washington

## Abstract

**Background:** Parkinson’s Disease (PD) affects millions globally, causing motor function impairments. Early detection is vital, and diverse data sources aid diagnosis. We focus on lower arm movements during keyboard and trackpad/touchscreen interactions, which serve as reliable indicators of PD. Previous works explore keyboard tapping and unstructured device monitoring, and we attempt to further these works with our structured tests taking account 2D hand movement in addition to finger tapping. Our feasibility study utilizes keystroke and mouse movement data from a structured online test conducted remotely combined with self-reported PD status to create a predictive model for detecting PD presence.

**Objective:** Through analysis of finger tapping speed and accuracy through keyboard input and 2-dimensional hand movement through mouse input, we differentiate between PD and non-PD participants. This comparative analysis enables us to establish clear distinctions between the two groups and explore the feasibility of using motor behavior to predict the presence of the disease.

**Methods:** Participants were recruited via email by the Hawaii Parkinson’s Association (HPA) and directed to a web application for the tests. The 2023 HPA symposium was also used as a forum to recruit participants and spread information about our study. The application recorded participant demographics, including age, gender, and race, as well as PD status. We conducted a series of tests to assess finger tapping, using on-screen prompts to request key presses of constant and random keys. Response times, accuracy, and unintended movements resulting in accidental presses were recorded. Participants performed a hand movement test consisting of tracing straight and curved on-screen ribbons using a trackpad or mouse, allowing us to evaluate stability and precision of two-dimensional hand movement. From this tracing, the test collected and stored insights concerning lower arm motor movement.

**Results:** Our formative study included 31 participants, 18 without PD and 13 with PD, and analyzed their lower limb movement data collected from keyboards and computer mice. From the dataset, we extracted 28 features and evaluated their significances using an ExtraTreeClassifier predictor. A Random Forest model was trained using the six most important features identified by the predictor. These selected features included insights into precision and movement speed derived from keyboard tapping and mouse tracing tests. This final model achieved an average F1-score of 0.7311 (±0.1663) and an average accuracy of 0.7429 (±0.1400) over 20 runs for predicting the presence of PD.

**Conclusion:** This preliminary feasibility study suggests the possibility of utilizing technology-based limb movement data to predict the presence of PD, demonstrating the practicality of implementing this approach in a cost-effective and accessible manner. In addition, this study demonstrates that structured mouse movement tests can be used in combination with finger tapping to detect PD.

## Introduction

In the United States alone, PD affects over one million individuals, with approximately 90,000 new diagnoses each year [1]. PD manifests with motor and non-motor symptoms that impact the entire body, including challenges like micrographia that significantly disrupt daily life [2] [3] [4] [5] [6]. Unfortunately, symptomatic evaluation remains the sole diagnostic method for PD, lacking an official diagnostic procedure. As a result, many cases go undiagnosed or misdiagnosed, hindering effective treatment [7] [8] [9] [10]. Moreover, even unofficial PD diagnostic tests are costly, requiring specialized equipment and laboratory procedures [11] [12] [13] [14]. Thus, there is an urgent need for scalable and accessible tools for PD detection and screening. Early diagnosis, which includes initiation of treatment and medication at an appropriate time, offers several benefits, including timely interventions and appropriate medication, leading to improved quality of life for patients [15] [16].

PD affects limb movements, particularly lower hand movements, as evidenced by multiple studies [5] [17] [18] [19] [20] [21]. Traditionally, clinical settings rely on neurologists who consider medical history, conduct physical examinations, and observe motor movements and non-motor symptoms for PD diagnosis [22] [23]. Recently, researchers have explored the use of smartphones as a measurement tool for PD detection [24] [25] [26]. Previous studies have shown that PD can be detected by monitoring digital device activity, such as abnormal mouse movements and atypical typing patterns [27] [28] [29] [30] [31] [32] [33]. Building on these findings, our goal is to develop a user-friendly web application that offers a cost-effective and accessible diagnostic method, overcoming the limitations of in-person examinations and smartphone tests.

Previous studies investigating the use of finger movement for PD detection often faced challenges in accessibility due to their requirements of specialized equipment like accelerometers and gyroscopes [34] [35]. For instance, Sieberts et al. utilized wearable sensors to gather accelerometer and gyroscope data, which might not be easily accessible to the general population [31]. Chandrabhatla et al. discussed the transition from lab-based to remote digital PD data collection, still relying on specific in-lab tools, which limited accessibility [36]. Skaramagkas et al. used wearable sensors to distinguish tremors, while Schneider et al. found PD distinctions using shoulder shrugs, arm swings, tremors, and finger taps [37] [38]. Their findings emphasized arm swings and individual finger tremors as significant indicators.

Numerous studies have leveraged mobile applications, like a work by Deng et al., which employed the Mpower app to assess movement [32] [33]. It is worth noting that older individuals, who are more vulnerable to PD, might not be as familiar with handheld devices like phones and tablets compared to computers and laptops, which are more common among them [39] [40] [41] [42]. Mobile phones became widely used in the early 2000s, with smartphones gaining popularity later, making them less familiar to the elderly [43] [44] [45]. On the other hand, many older adults have more experience with computers, which have been in use for a longer time [46]. This familiarity not only expands the potential participant group but also ensures more reliable data collection due to participants’ better understanding of the test procedures [47] [48] [49].

Keyboard typing’s potential for PD prediction has also been explored. In a study by Arroyo-Gallego et al., the neuroQWERTY method was used, employing computer algorithms that consider keystroke timing and subtle movements to detect PD [50]. This approach was extended to uncontrolled at-home monitoring, using participants’ natural typing and laptop interaction to detect signs of PD. The algorithm’s performance at-home nearly matched its in-clinic efficacy. However, the lack of structure in this approach makes direct performance comparisons challenging. Additionally, Noyce et al. investigated genetic mutations and keyboard tapping over 3 years [51]. They calculated risk scores using PD risk factors and early features. However, this study involves genetic information, which might not be accessible to many patients.

While drawing tests have been well-studied, the investigation of using mouse hovering to trace specific paths is limited. Unlike freehand tablet drawing, which is flexible but lacks regulated data, Isenkul and Sakar used a tablet to help PD patients with micrographia [52]. Yet, since touchscreens are scarce on larger devices, a mouse provides a more accessible comparison [53] [54].

Taking a different approach, researchers have used brain scans and biopsies to study changes in PD-related brain regions. Kordower et al. observed the progression of nigrostriatal degradation in PD patients over time. By analyzing brain regions, they found that the loss of dopamine markers four years after diagnosis indicated PD [55].

These prior studies collectively contribute valuable insights into using digital devices for gathering motor-related PD data. Our research aims to expand upon this inspirational prior work by exploring the potential of an easily accessible online test involving keyboard finger tapping and mouse movements. We use a web application compatible with common devices to analyze these data, distinguishing individuals with and without PD, to assess the feasibility of a more accessible and cost-effective detection method.

While in-lab devices are precise but less accessible due to cost, web-based tests are cost-effective and accessible. Mobile apps are user-friendly but less familiar to older PD patients. Our web app, accessible on various devices, particularly computers, ensures familiarity and consistency for reliable data. Our method furthers freeform drawings and keyboard tapping with structured tracing and key tapping tests, allowing direct performance comparisons.

We present an affordable and accessible method for PD detection using a web application that captures and analyzes lower hand movements during keyboard and mouse interactions. Keystrokes are measured by logging the time interval between prompts and keypresses, while false presses are recorded to detect finger shaking. Mouse movement is tracked every 500 milliseconds to assess precision and identify shaking or unintended movements. In a remote study with 31 participants, including 13 PD patients and 18 non-PD controls, we trained a machine learning model on six extracted movement features, achieving promising predictive performance. The model yielded an average F1 score of 0.7311 and an average accuracy of 0.7429. These results demonstrate the practicality of technology-driven limb movement data collection for effective PD detection.

## Methods

### IRB Approval

This study obtained approval from the UH Manoa Institutional Review Board (protocol #2022-00857). Ensuring accurate identification of PD among participants was a significant challenge due to the lack of an official diagnostic certificate for PD. We therefore relied on self-report, and we required users to confirm their results with an intrusive dialogue that had to be dismissed before the test commenced, minimizing mistakes.

### Recruitment

Participants were recruited through the HPA and similar organizations. We collaborated with the former president of the HPA, who shared detailed information about our test via email. We set up a booth at the 2023 Hawai’i Parkinson’s Symposium, an event coordinated by the HPA. Attendees had the chance to take the test using a provided device and receive slips with the test URL. We welcomed participation from both PD and non-PD individuals who showed interest. Participants recruited by email were provided with a web application link to conveniently complete the tests remotely. The study included individuals from both the PD and non-PD groups. This feasibility study consisted of a cohort of 31 participants. The age distribution was 65.226 +/- 10.832 years for all participants, 69 +/- 7.147 years for PD participants, and 62.5 +/- 12.144 for non-PD participants.

Recognizing the limitations of our small sample size, we stress that this study represents an initial investigation into the utility of this test for PD detection. Our plan is to build upon these results through a larger-scale study involving a broader range of participants.

To address potential misclassification associated with using self-reporting, we implemented a comprehensive strategy. Participants were presented with an intrusive dialog box containing their entered demographics, including PD status, and were required to review and confirm its accuracy. They had the flexibility to modify their status and demographics, minimizing errors. Demographic data collection followed IRB guidelines with support from the HPA. Participants were provided the option to select “prefer not to answer” for certain demographic questions, encouraging test completion even without specific demographic details, as they were not essential to the study’s objectives.

### Parkinson’s Test

Participants were instructed to type on a keyboard while the test recorded timestamps and finger movements corresponding to key positions (Figure 1). They pressed a specific key in response to on-screen signals, and we collected data on the expected key, pressed key, and response time. The measurement process had three levels of increasing difficulty with greater key randomization. The lowest difficulty of keyboard tapping prompted the press of a single key ten times, while the second level alternated between two keys for ten trials. The third difficulty changed the requested key to a random one for every press of ten trials. For trackpad/mouse data, participants hovered the mouse along a designated path. This path was created in a way such that it took up certain percentages of the screen, as opposed to a set number of pixels, enabling it to adapt to the screen of the device being used and present an equal test to all participants. Starting with a straight line, subsequent levels introduced a sine wave-like shape and a spiral shape. Participants could monitor their progress by observing a highlighted portion of the shape, guided by animated direction indicators and “start”/“finish” markings. The interface of the web application, including those for demographic data collection and mouse and keyboard test administration, is show in Figure 2. We recorded data on position, time, and whether the mouse was inside or outside the indicated area. We also recorded the height and width of the participant’s device so it could be considered for calculations and could be used to recreate the user’s test.

**Figure 1.**
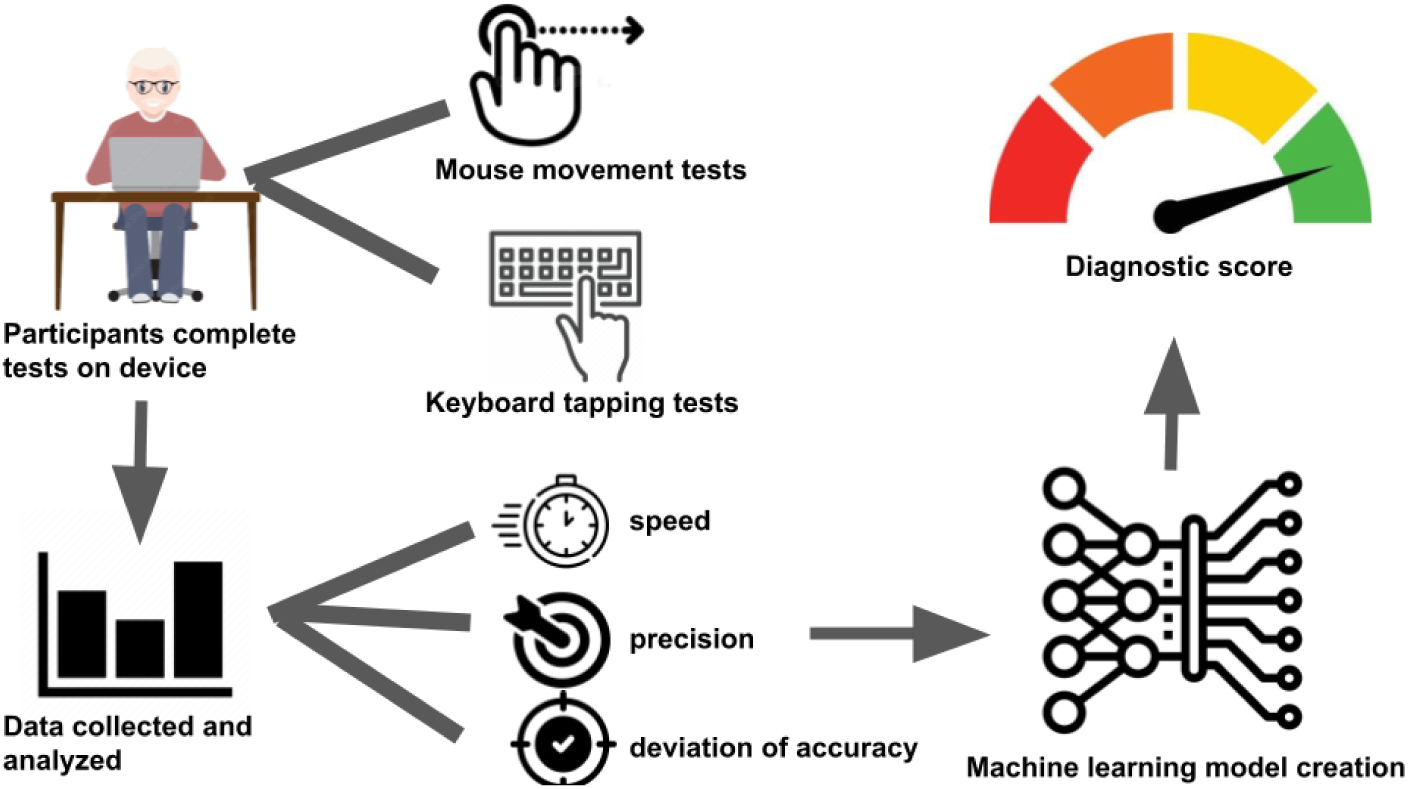
Overall study design. Participants completed mouse movement and keyboard tapping tests on their device, from which data was collected and analyzed for speed, precision, and accuracy. A machine learning model was trained on these data to predict the presence of PD.

**Figure 2.**
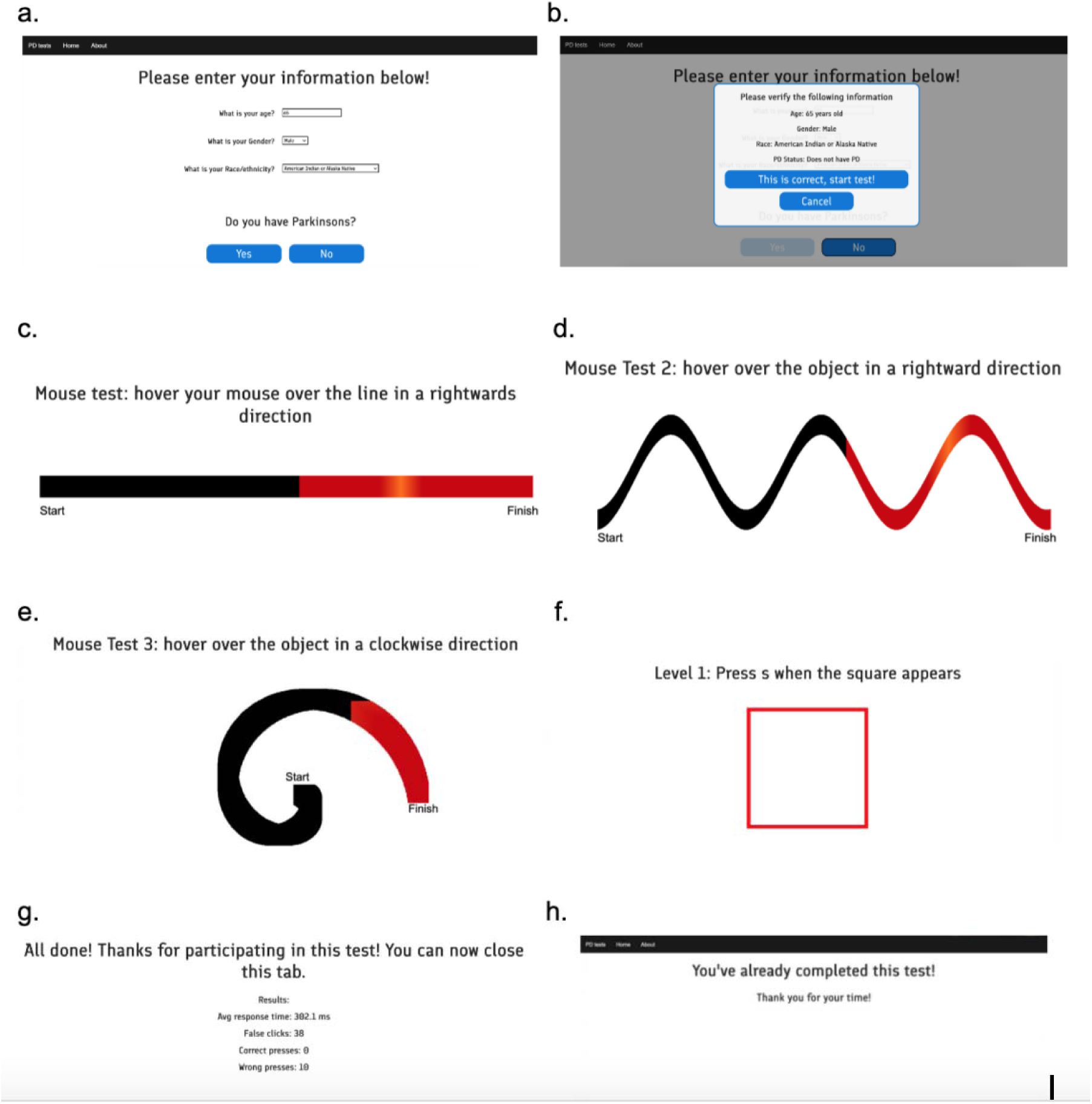
Screenshots of the web application: a) Information is collected through an online form, b) The participant is asked to confirm the entered information for correctness to prevent mistakes, c) The linear mouse test asks users to trace the ribbon in a leftward direction, d) The sine-wave mouse test asks users to trace through the region, e) The spiral mouse test asks users to trace a spiral, f) The keyboard test asks users to press a certain letter when the red square prompt appears, g) After completion, the test informs the participant and thanks them for their time, h) The test informs the user that they have already completed the test after completion, minimizing duplicate entries.

We used a custom web application created with HTML, JavaScript, and CSS to collect data. For keyboard tapping, an HTML canvas was used to display a red square as a prompt. JavaScript was used to tracked keypress timing and calculated reaction times. For mouse tracing, an HTML canvas produced visuals of straight lines, sine waves, and spirals with direction indicator animations. JavaScript determined cursor location within the designated area and recorded mouse coordinates every 500 milliseconds.

After completing both tests, the collected data was securely transmitted and stored using a deta.sh base facilitated by the deta.sh micros API, ensuring efficient data management.

This test was taken by both PD and non-PD participants primarily aged between 50 and 80 years (Figure 3). The male/female ratio was nearly equal (Figure 4), and the race of participants was predominantly White and Asian (Figure 5). In addition, the PD vs non-PD participant ratio was slightly skewed towards non-PD patients (Table 1).

**Figure 3.**
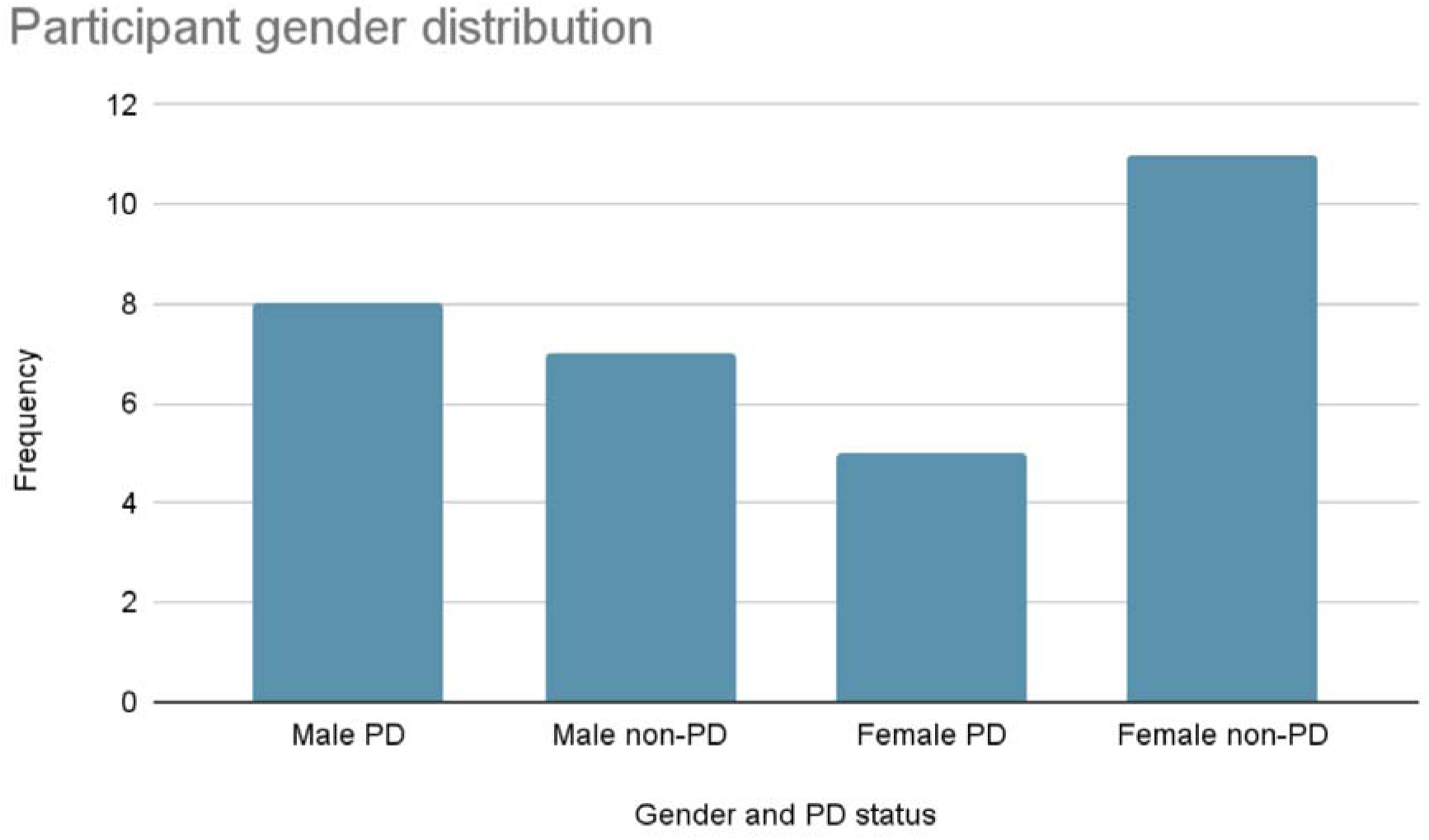
Participant age distribution: The average age for all participants was 65.226 years with a standard deviation of 10.832. For PD participants, the average age was 69 years with a standard deviation of 7.147. For non-PD participants, the average age was 62.5 years with a standard deviation of 12.144.

**Figure 4.**
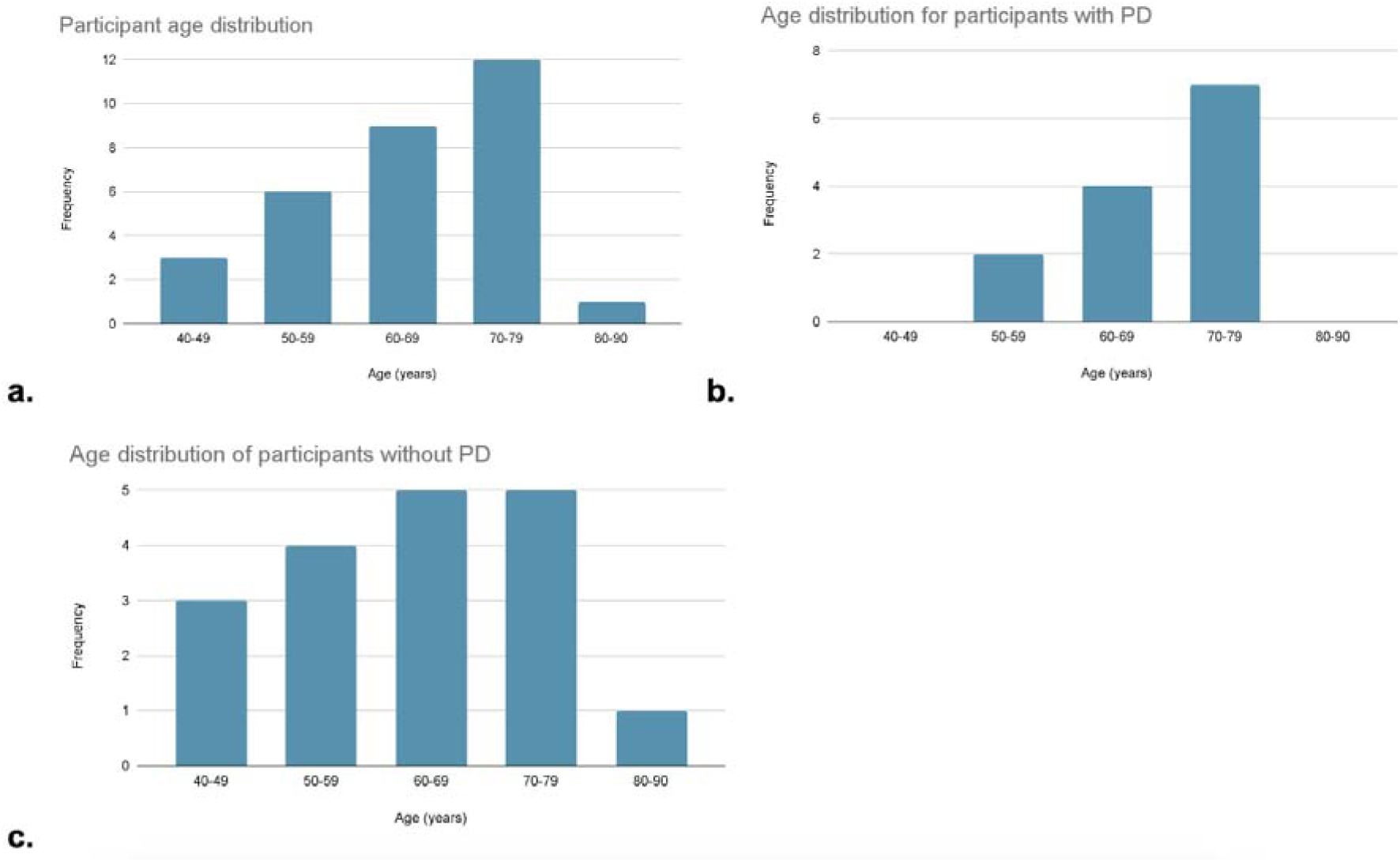
Gender distribution: Mostly balanced with a 15:16 Male: Female ratio, with the total distribution slightly skewed towards female non-PD participants.

**Figure 5:**
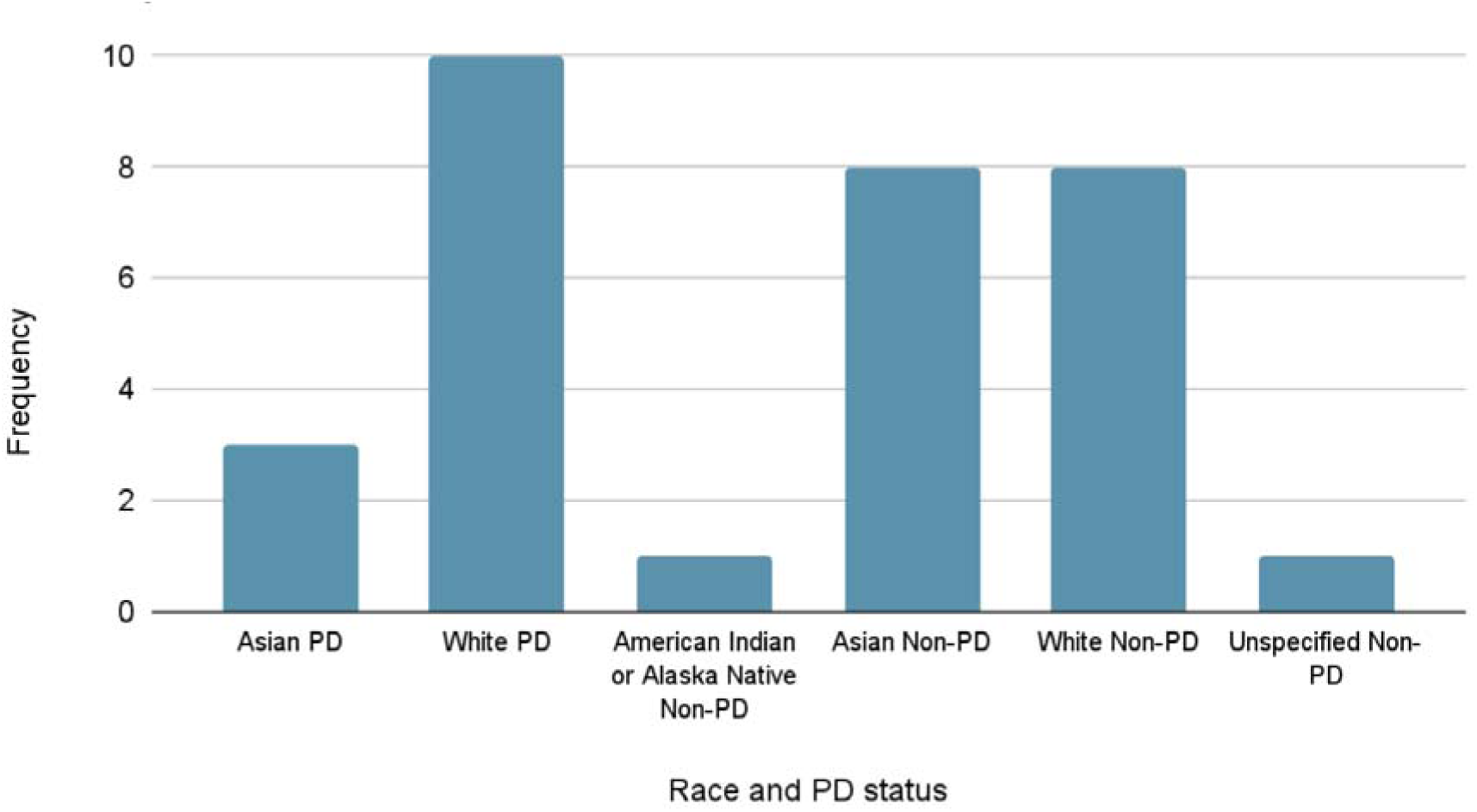
Race distribution: Largely skewed towards White and Asian Participants, with Whites, American Indian/Alaska Natives having a similar amount of PD and non-PD participants, with Asian participants mostly being non-PD.

**Table 1.**
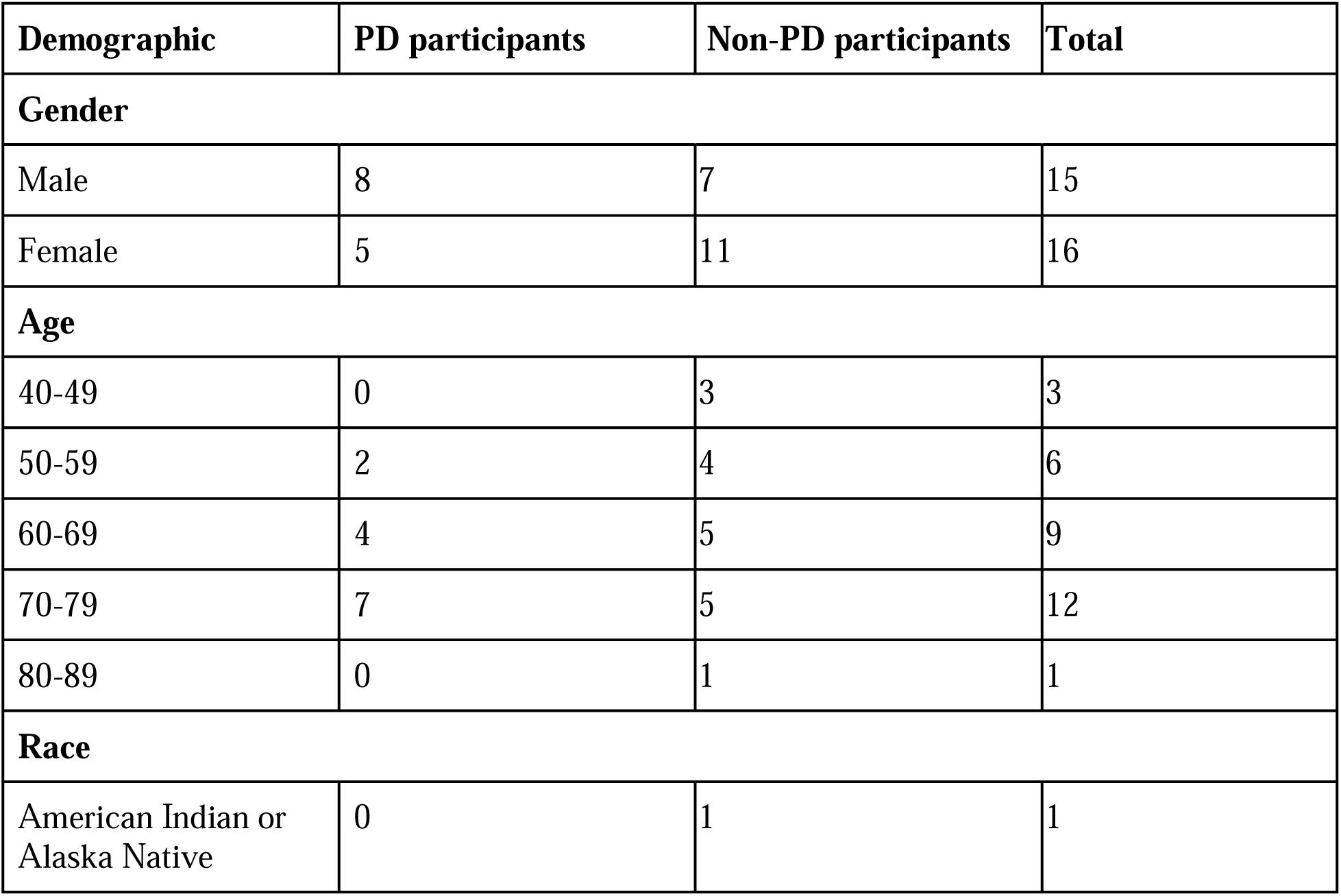
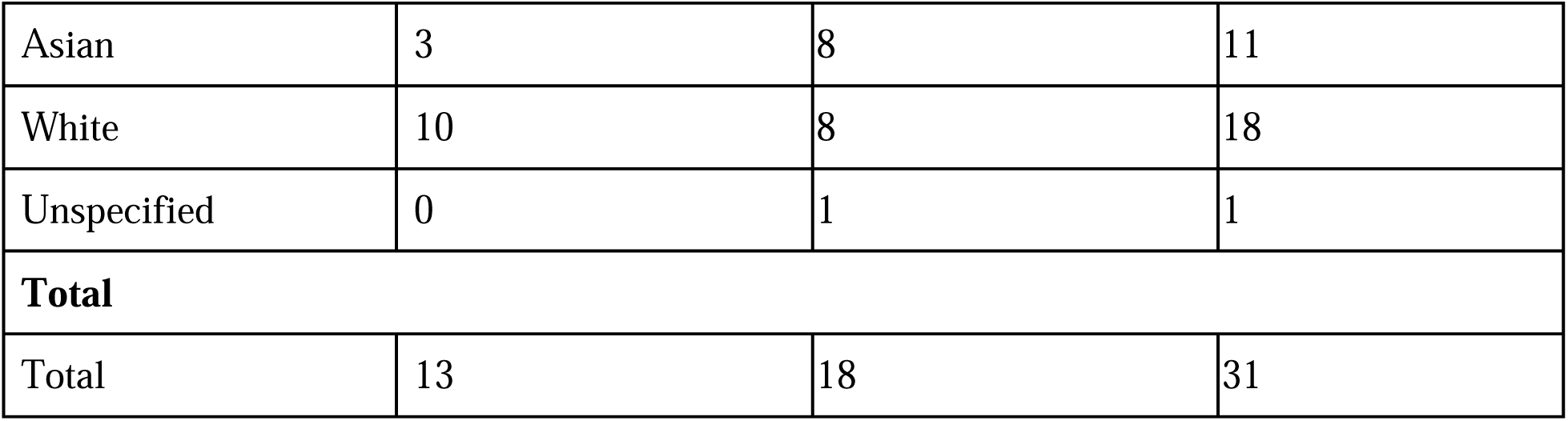
Participant Demographics. Displays the gender, age, and race distributions for PD, non-PD, and all participants.

| Demographic | PD participants | Non-PD participants | Total |
| --- | --- | --- | --- |
| **Gender** |  |  |  |
| Male | 8 | 7 | 15 |
| Female | 5 | 11 | 16 |
| **Age** |  |  |  |
| 40-49 | 0 | 3 | 3 |
| 50-59 | 2 | 4 | 6 |
| 60-69 | 4 | 5 | 9 |
| 70-79 | 7 | 5 | 12 |
| 80-89 | 0 | 1 | 1 |
| **Race** |  |  |  |
| American Indian or Alaska Native | 0 | 1 | 1 |
| Asian | 3 | 8 | 11 |
| White | 10 | 8 | 18 |
| Unspecified | 0 | 1 | 1 |
| **Total** |  |  |  |
| Total | 13 | 18 | 31 |

### Statistical Analysis

We conducted a series of tests to measure keyboard keypresses, false presses, and timestamps as proxies for unintended movements, shaking, and anomalies. Through mouse hovering tests, we observed continuous mouse movement controlled by participants’ arms and recorded deviations from the centerline. These tests provided insights into events such as accidental deviations or vibrations, potentially indicative of PD symptoms. Additionally, we examined mouse data, focusing on hovering speed and precision as indicators of unintentional lower arm movements. We observed differences between participants with and without PD that could serve as potential disease indicators. These findings contribute to the advancement of PD detection methods by leveraging discernible distinctions between individuals with and without the condition.

During the test, we collected a total of 17 features in four major categories. One category assessed participant hand stability while tracing a straight line, considering mouse coordinates (x and y), deviation from the centerline, and whether the mouse was inside the given region or not were measured every 500 milliseconds. Another category focused on tracing a curved line while maintaining hand stability, for which the coordinates (x and y) and whether the mouse was inside the region or not was recorded. The third category measured response times and accuracy of key presses prompted by visual cues to evaluate reaction speed. The final category recorded the number of false presses during keyboard prompts as an indicator of unintended hand movements.

We extracted 28 features from each participant’s test. These included 17 baseline features along with additional features derived as a function of the baseline features. For instance, analyzing tracing deviation from the centerline produced multiple features like mean and max deviations. Incorporating screen width into time taken for tracing improved the feature’s association with PD, as seen in Table 2. An ExtraTreeClassifier predictor assessed the importance of each feature. We identified six key features as the most indicative of PD. These features were used to train a Random Forest model. These six features were selected due to their significantly higher importance scores compared to other related features as reported by the predictor. The selected features included mean deviation during straight line tracing, time to trace sine wave relative to window width, spiral tracing time, average false presses, total response time for constant key tapping, and accuracy in responding to random key prompts.

**Table 2.** Feature enhancement. A sample of features that were improved by considering other aspects of the data that affected them.

| Feature | Enhancement | Original model F1 score | Model F1 score for enhanced feature | F1 score improvement |
| --- | --- | --- | --- | --- |
| Amount of time taken for tracing straight line | Feature taken with respect to window width | 0.6528 +/- 0.1960 | 0.7167 +/- 0.1633 | 0.0639 |
| Number of correctly pressed keys when prompted with a random key | Feature taken with respect to average keypress time | 0.6333 +/- 0.1944 | 0.6944 +/- 0.2641 | 0.0611 |

Our Random Forest model was trained using an 80/20 train/test split. Due to the internal out-of-bag evaluation of Random Forest models, a separate validation set was not used. The model underwent 20 rounds of training with new train/test splits using an 80/20 ratio before for each round. We used this evaluation method to account for our dataset’s small size.

## Results

Our Random Forest machine learning (ML) model, trained on six features involving line tracing, sinusoid tracing, spiral tracing, accuracy and speed of keypress prompts, and false presses, yielded an average F1 score of 0.7311 with a standard deviation of 0.1663. It also achieved an average accuracy of 0.7429 with a standard deviation of 0.1400 (Table 3). All measurements were taken over 20 independent runs, with randomly sampled train/test splits created in each run. In addition, we trained the same set of 6 high-performing features on different types of ML models determine the optimal model type (Table 4).

**Table 3:**
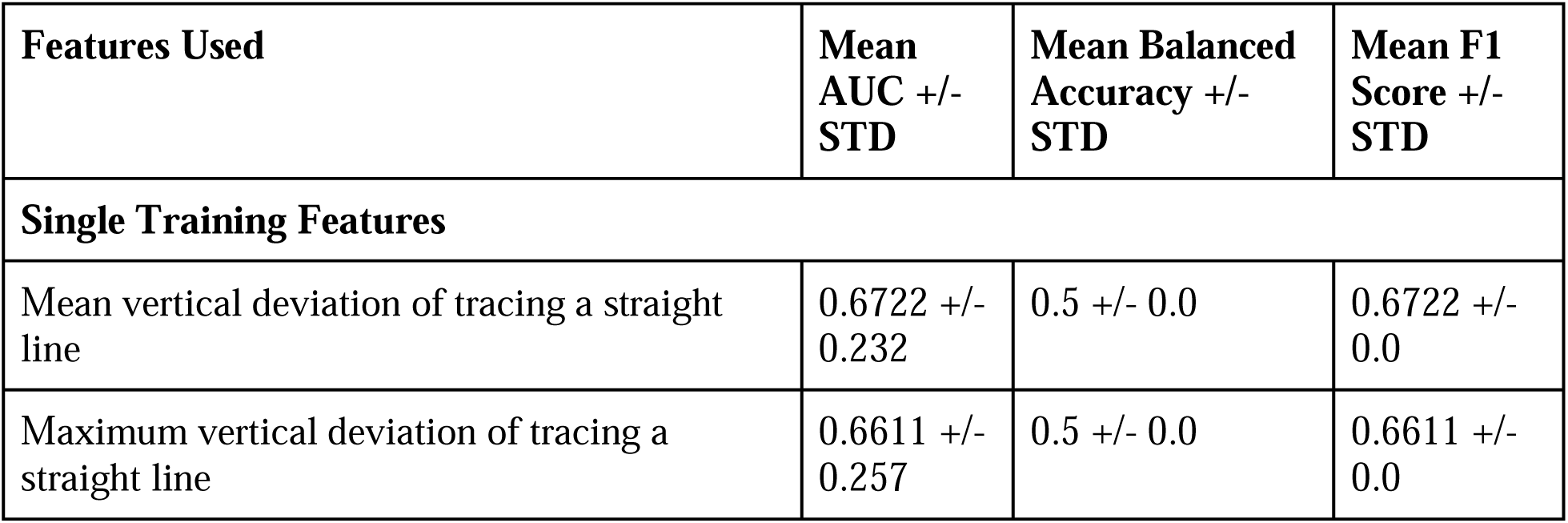
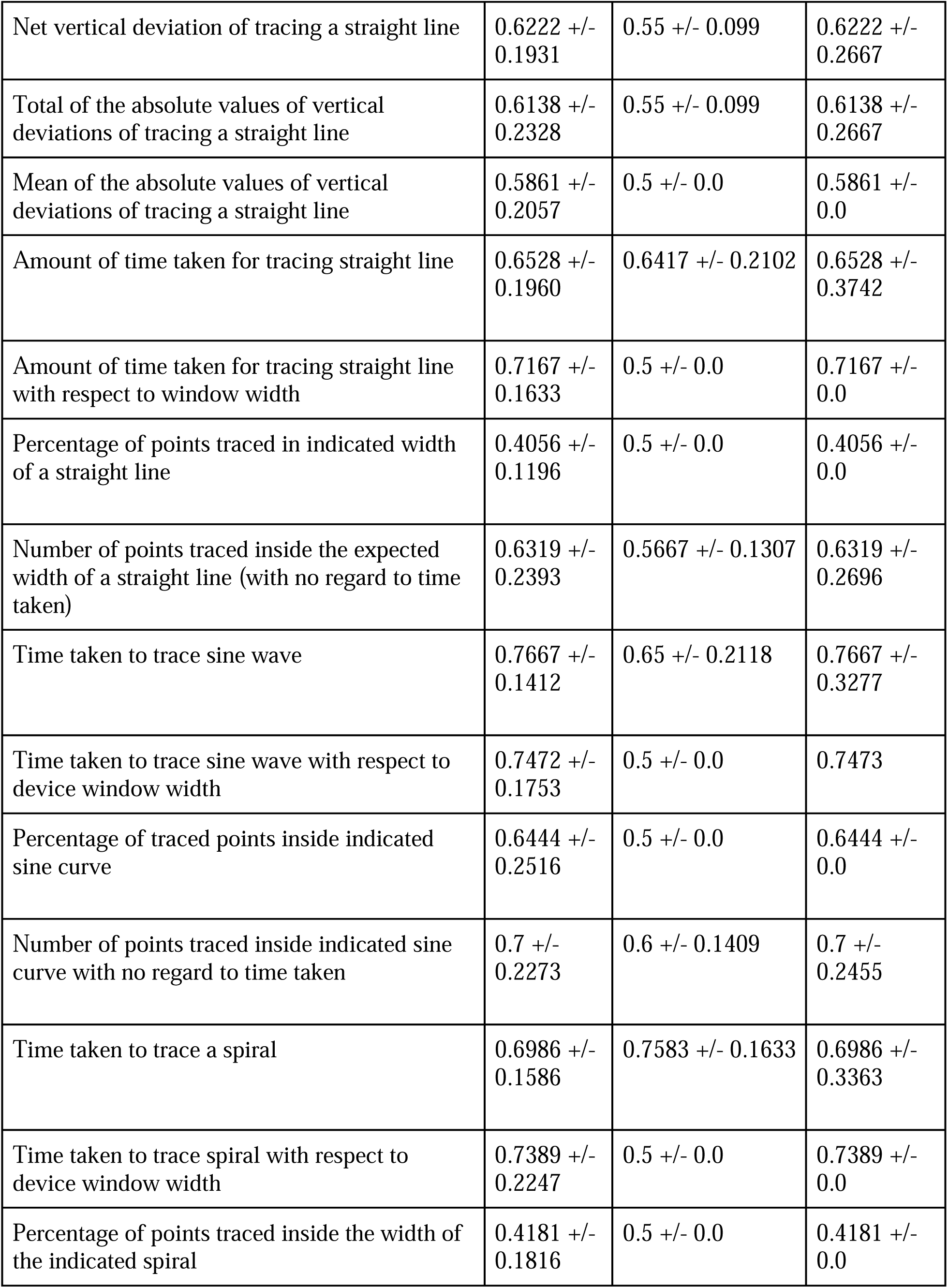
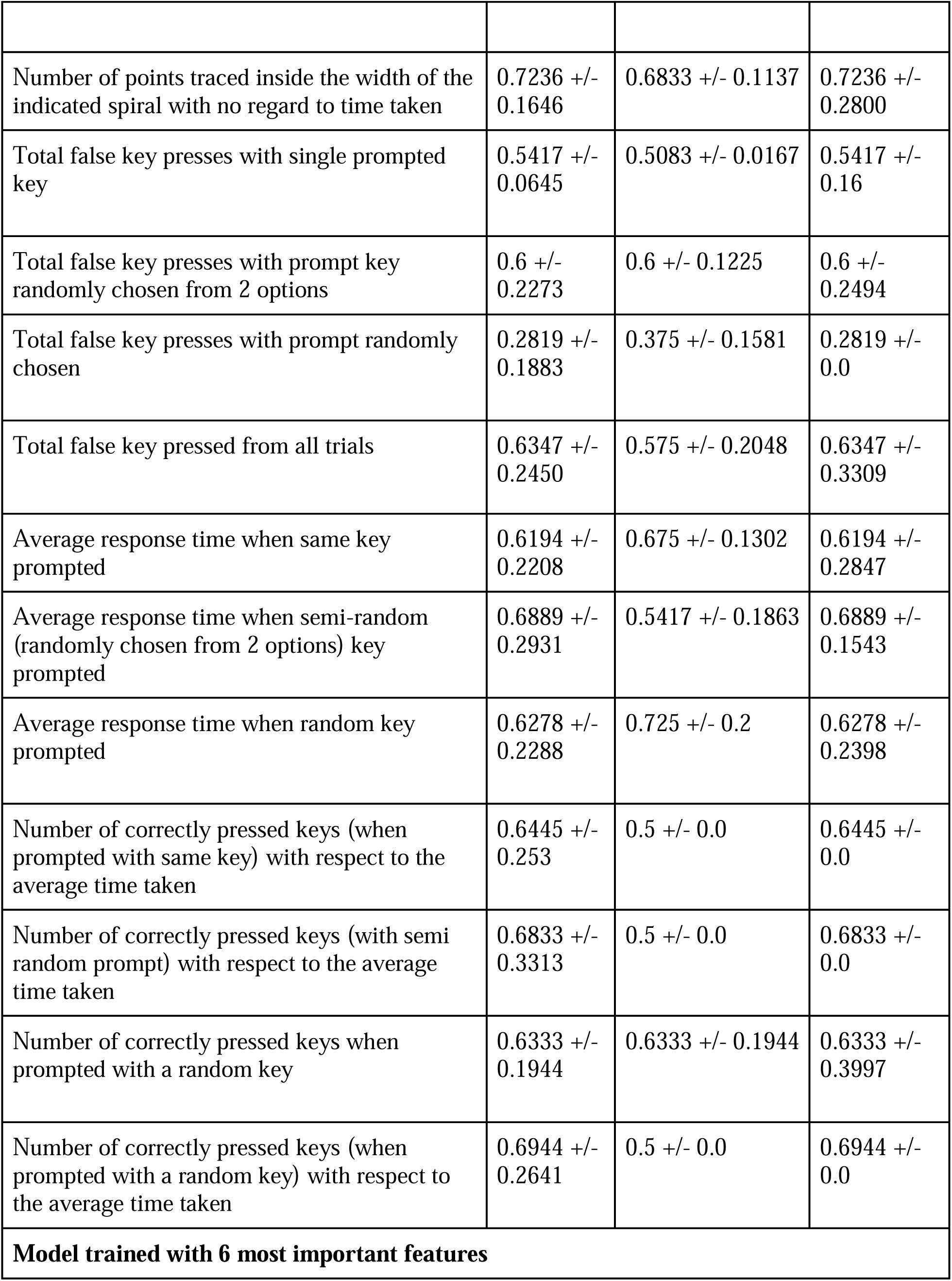
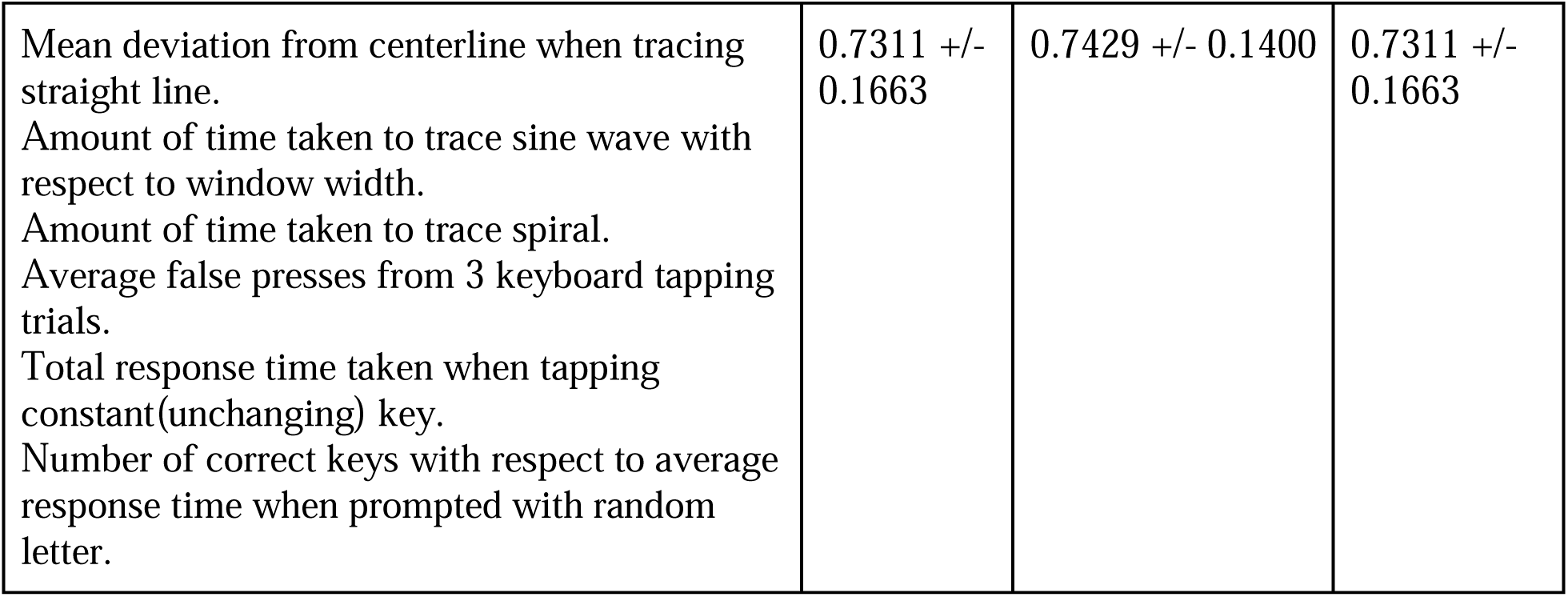
Mean AUC, Balanced Accuracy, and F1 score of models trained on single and multiple features. Each individual feature was trained to evaluate its efficacy, shown in the first 28 content rows. An ExtraTreeClassifier was used to rank features by importance. The 6 most important features were used to train a final Random Forest model, shown in the last row.

| Features Used | Mean AUC +/- STD | Mean Balanced Accuracy +/- STD | Mean F1 Score +/- STD |
| --- | --- | --- | --- |
| **Single Training Features** |  |  |  |
| Mean vertical deviation of tracing a straight line | 0.6722 +/- 0.232 | 0.5 +/- 0.0 | 0.6722 +/- 0.0 |
| Maximum vertical deviation of tracing a straight line | 0.6611 +/- 0.257 | 0.5 +/- 0.0 | 0.6611 +/- 0.0 |
| Net vertical deviation of tracing a straight line | 0.6222 +/- 0.1931 | 0.55 +/- 0.099 | 0.6222 +/- 0.2667 |
| Total of the absolute values of vertical deviations of tracing a straight line | 0.6138 +/- 0.2328 | 0.55 +/- 0.099 | 0.6138 +/- 0.2667 |
| Mean of the absolute values of vertical deviations of tracing a straight line | 0.5861 +/- 0.2057 | 0.5 +/- 0.0 | 0.5861 +/- 0.0 |
| Amount of time taken for tracing straight line | 0.6528 +/- 0.1960 | 0.6417 +/- 0.2102 | 0.6528 +/- 0.3742 |
| Amount of time taken for tracing straight line with respect to window width | 0.7167 +/- 0.1633 | 0.5 +/- 0.0 | 0.7167 +/- 0.0 |
| Percentage of points traced in indicated width of a straight line | 0.4056 +/- 0.1196 | 0.5 +/- 0.0 | 0.4056 +/- 0.0 |
| Number of points traced inside the expected width of a straight line (with no regard to time taken) | 0.6319 +/- 0.2393 | 0.5667 +/- 0.1307 | 0.6319 +/- 0.2696 |
| Time taken to trace sine wave | 0.7667 +/- 0.1412 | 0.65 +/- 0.2118 | 0.7667 +/- 0.3277 |
| Time taken to trace sine wave with respect to device window width | 0.7472 +/- 0.1753 | 0.5 +/- 0.0 | 0.7473 |
| Percentage of traced points inside indicated sine curve | 0.6444 +/- 0.2516 | 0.5 +/- 0.0 | 0.6444 +/- 0.0 |
| Number of points traced inside indicated sine curve with no regard to time taken | 0.7 +/- 0.2273 | 0.6 +/- 0.1409 | 0.7 +/- 0.2455 |
| Time taken to trace a spiral | 0.6986 +/- 0.1586 | 0.7583 +/- 0.1633 | 0.6986 +/- 0.3363 |
| Time taken to trace spiral with respect to device window width | 0.7389 +/- 0.2247 | 0.5 +/- 0.0 | 0.7389 +/- 0.0 |
| Percentage of points traced inside the width of the indicated spiral | 0.4181 +/- 0.1816 | 0.5 +/- 0.0 | 0.4181 +/- 0.0 |
| Number of points traced inside the width of the indicated spiral with no regard to time taken | 0.7236 +/- 0.1646 | 0.6833 +/- 0.1137 | 0.7236 +/- 0.2800 |
| Total false key presses with single prompted key | 0.5417 +/- 0.0645 | 0.5083 +/- 0.0167 | 0.5417 +/- 0.16 |
| Total false key presses with prompt key randomly chosen from 2 options | 0.6 +/- 0.2273 | 0.6 +/- 0.1225 | 0.6 +/- 0.2494 |
| Total false key presses with prompt randomly chosen | 0.2819 +/- 0.1883 | 0.375 +/- 0.1581 | 0.2819 +/- 0.0 |
| Total false key pressed from all trials | 0.6347 +/- 0.2450 | 0.575 +/- 0.2048 | 0.6347 +/- 0.3309 |
| Average response time when same key prompted | 0.6194 +/- 0.2208 | 0.675 +/- 0.1302 | 0.6194 +/- 0.2847 |
| Average response time when semi-random (randomly chosen from 2 options) key prompted | 0.6889 +/- 0.2931 | 0.5417 +/- 0.1863 | 0.6889 +/- 0.1543 |
| Average response time when random key prompted | 0.6278 +/- 0.2288 | 0.725 +/- 0.2 | 0.6278 +/- 0.2398 |
| Number of correctly pressed keys (when prompted with same key) with respect to the average time taken | 0.6445 +/- 0.253 | 0.5 +/- 0.0 | 0.6445 +/- 0.0 |
| Number of correctly pressed keys (with semi random prompt) with respect to the average time taken | 0.6833 +/- 0.3313 | 0.5 +/- 0.0 | 0.6833 +/- 0.0 |
| Number of correctly pressed keys when prompted with a random key | 0.6333 +/- 0.1944 | 0.6333 +/- 0.1944 | 0.6333 +/- 0.3997 |
| Number of correctly pressed keys (when prompted with a random key) with respect to the average time taken | 0.6944 +/- 0.2641 | 0.5 +/- 0.0 | 0.6944 +/- 0.0 |
| **Model trained with 6 most important features** |  |  |  |
| Mean deviation from centerline when tracing straight line.<br>Amount of time taken to trace sine wave with respect to window width.<br>Amount of time taken to trace spiral.<br>Average false presses from 3 keyboard tapping trials.<br>Total response time taken when tapping constant(unchanging) key.<br>Number of correct keys with respect to average response time when prompted with random letter. | 0.7311 +/- 0.1663 | 0.7429 +/- 0.1400 | 0.7311 +/- 0.1663 |

**Table 4:** Mean AUC, Balanced Accuracy, and F1 score of different types of models trained. We trained several types of models on the same set of the 6 most important features and evaluated their average metrics over 20 runs. Between each run, the train/test split was resampled, maintaining the 80:20 ratio.

| Model trained (20 runs) | Accuracy +/- STD | F Score +/- STD |
| --- | --- | --- |
| Random Forest Classifier | 0.7429 +/- 0.1400 | 0.7311 +/- 0.1707 |
| Decision Tree Regression | 0.5999 +/- 0.1245 | 0.5862 +/- 0.1455 |
| Support Vector Classifier (SVC) | 0.6071 +/- 0.1268 | 0.5816 +/- 0.1622 |
| MLP Classifier | 0.5214 +/- 0.1707 | 0.3740 +/- 0.1939 |

We identified high-performing features and further analyzed their relationship with participants’ body movements. By reconstructing the traces based on the collected data, visual differences between PD and non-PD participants were observed. As shown in Figure 6, while most straight-line traces showed similarities, PD participant traces exhibited sudden irregularities, whereas non-PD traces contained minimal irregularities. Additionally, significant differences were observed in sinusoid traces, with PD participants completing the test faster but with more irregularities and fewer points traced within the designated area. Conversely, non-PD participants took more time but demonstrated greater precision. Similar patterns emerged in spiral traces, where PD participants traced the spiral more rapidly but with less precision compared to non-PD participants.

**Figure 6:**
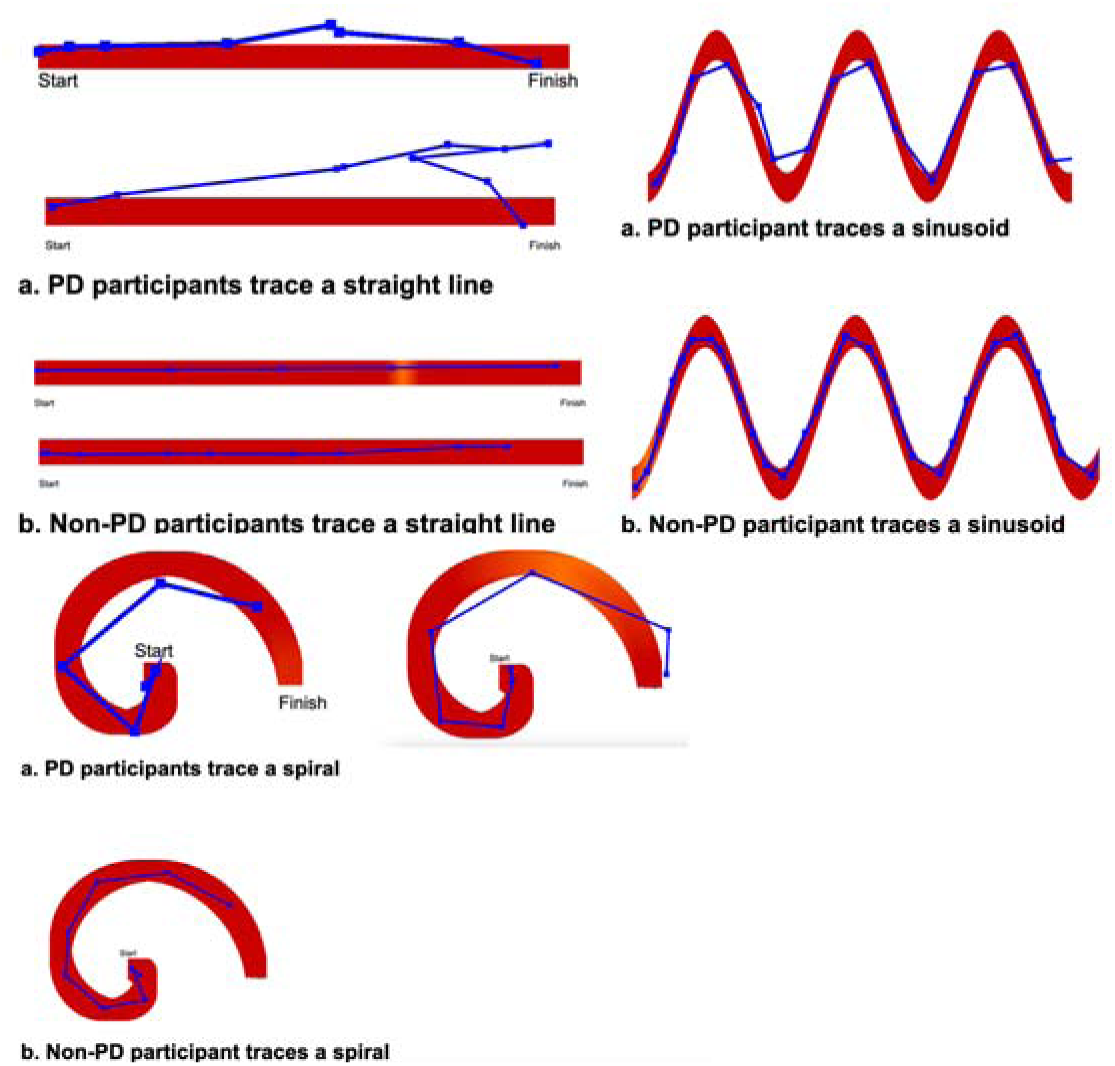
Sample traces of PD and non-PD participants. Generally, PD participant traces can be observed to be more irregular and less precise than those of non-PD participants.

The performance of this model supports the feasibility of the automatic detection of PD through hand and finger movement analysis. These findings support the feasibility of using traced lines and curves as a potential method for predicting the presence of PD and other related conditions affecting limb movements using ubiquitous consumer devices such as a laptop.

## Discussion

### Principal Results

This study provides evidence supporting the feasibility of remote collection of limb movement data using ubiquitously available consumer technology. We addressed concerns regarding device variations by considering device performance and specifications, such as screen height and width, which were measured and recorded by the application. Interestingly, excluding the time taken to complete the test improved the results for many extracted features. The findings suggest that either device performance affected the timing data or PD has limited influence on hand movement speed indicators such as key tapping or drawing. However, the latter scenario is unlikely, as multiple studies have shown that PD affects key tapping speed [56, 57]. Additionally, false presses when prompted with a random key do not seem to be linked to PD, as this feature’s performance is notably weak. This might be due to displaying the next key before the prompt, leading to unintended key presses due to hand repositioning, a phenomenon common in older individuals regardless of PD status. Furthermore, our findings reinforce the use of measuring limb movements as an indicator of PD presence. The majority of models trained on individual extracted features yielded mean F1 scores and AUC values surpassing 0.5, indicating a weak but existing correlation between the feature and the existence of PD. Additionally, 82.14% (23/28) of the features achieved an F1 score of at least 0.6, while 21.4% (6/28) achieved an F1 score of at least 0.7. Our final and most optimal model was able to achieve an accuracy of 74.29% and an F1 score of 73.11% These results highlight a clear correlation between the speed and precision of tracing movements and the speed and accuracy of finger tapping with the presence of PD.

### Comparison to Previous Work

This study extends prior research by bringing lab-based movement testing to remote assessment on personal devices, enhancing accessibility and scalability. Unlike studies monitoring regular keyboard use, we employ a structured test for better comparability. Additionally, our app includes structured tracing tests, exploring other motor aspects beyond keyboard tapping as a PD indicator. Our model’s performance is comparable to the at-home neuroQWERTY test by Teresa et al., which acheived an Area Under Curve (AUC) of 0.7311, while the clinic-based neuroQWERTY test achieves 0.76 AUC [58]. However, our model’s accuracy lags behind clinical tests with a similar aim, like Tsoulos, Ioannis G., et al., achieving 93.11% PD detection accuracy, and Memedi, Mevludin, et al., reaching 84% PD detection accuracy [59, 60].

### Limitations

A key limitation of this study concerns device consistency. Using a specific device may introduce biases due to user unfamiliarity, while allowing participants to use their own devices may result in performance and specification variations. To address this, we standardized the collected data and recorded device aspects such as display width, height, and frames per second. This information enabled us to assess the influence of screen dimensions and device performance on user results. For example, by comparing the user’s mouse coordinates to the screen width in pixels, we determined the percentage of the screen that the mouse had moved, rather than the raw amount of pixels, which would depend on the device used. However, other factors affecting the collected data, including differences between trackpads and mice as well as keyboard types have not been accounted for. These differences may have influenced our collected data and impacted our results. Additionally, the remote nature of the study posed a challenge as participants completed it without supervision, potentially introducing errors and impacting results. It is worth noting that the PD-to-non-PD participant ratio was 13:18, leading to an imbalance that could affect data analysis. Moreover, the study predominantly included participants of White and Asian ethnicities, introducing a racial imbalance that may impact the model’s accuracy for other racial groups if race influences the final prediction.

An additional limitation concerns the age difference in our sample. The non-PD participants had an average age of 62.5, while PD participants had an average age of 69. Since we did not adjust for this age disparity, it may have influenced our results. In addition, the lack of an official PD diagnosis led us to rely solely on self-reports. Despite efforts to enhance accuracy, errors could have affected results. Furthermore, it is recognized that conditions like essential tremor (ET) cause symptoms similar to PD, potentially leading to misdiagnoses [61]. This scenario might have skewed our findings if individuals diagnosed with PD had ET. This concern could be addressed by inquiring about participants’ history of ET prior to the test and taking this into account for analysis.

### Further research

Machine learning (ML) holds promise for predicting movement-related conditions, including essential tremor, and its application can be extended to other movement-impacting diseases. Standardizing a comprehensive test could offer individuals a single, straightforward assessment to evaluate their likelihood of having different health conditions. By incorporating diverse shapes for tracing, such as those involving sudden stops or changes in direction, additional valuable insights into hand movements of both PD and non-PD participants can be gleaned. To address limitations associated with unsupervised remote studies, a supervised approach with real-time monitoring could be implemented, providing immediate feedback to ensure protocol adherence and improve data reliability. Additionally, collecting information on participants’ device types can help address potential bias arising from device disparities. With a sufficiently large sample size, subgroup analysis based on device type could mitigate the impact of device variations on data and strengthen the validity of the findings. Some related studies have used significantly more participants [59, 60]. Expanding the participant sample size would support the generalizability of our findings.

Another potentially fruitful avenue of expanding PD screening tools would be to include additional data modalities such as computer vision. Computer vision analysis has been successfully employed for a variety of health screening and diagnostic tasks [62] [63] [64] [65] [66], including abnormal hand movements [67] and movement of other body parts [68] for conditions such as autism. Utilizing such techniques for PD screening can expand the performance of the tools through a more comprehensive and multimodal analysis.

## Acknowledgements

We gratefully acknowledge the Hawaii Parkinson’s Association for leading our recruitment efforts and informing us about the need for better screening tools for Parkinson’s Disease.

## Data Availability

The data collected from our study has been de-identified and is publicly available at https://github.com/skparab2/Parkinsons_movement_tests_dataset.

## Abbreviations

PD: Parkinson’s Disease

